# A deterministic framework for linkage of infectious disease surveillance and national mortality records: development and validation using tuberculosis surveillance data

**DOI:** 10.64898/2026.11.12.26367852

**Authors:** Orla Presslie, Ailva O’Reilly, Dominik Zenner, Jamilah Meghji, Manish Pareek, Sharon E Cox

## Abstract

**Background:** Linkage of infectious-disease surveillance data to mortality records is important for estimating long-term outcomes, but incomplete personal identifiers present methodological challenges. We developed and validated a deterministic linkage framework for linking the National Tuberculosis Surveillance System (NTBS) with Office for National Statistics (ONS) mortality records.

**Methods:** TB notifications in England from 2001–2024 were linked to ONS mortality records using 12 sequential matching rules based on combinations of NHS number, date of birth, name, postcode, sex and phonetic matching. Linkage outputs underwent person-level deduplication and clerical review. Precision and recall were estimated, and potential linkage bias assessed by variation in identifier completeness and linkage success. Confirmed false-positive links identified during validation were removed from the analytical dataset.

**Results:** Among 151,965 TB notifications, 28,060 initial links were identified. Following deduplication, validation and refinement, 26,937 linked notifications were retained. Estimated precision increased from 97.7% to 99.2% (95% CI 96.8–100.0) following removal of false-positive links; estimated recall was 98.1% (95% CI 95.4–98.6%). More than 90% of links used NHS-number-based match keys. PDS enrichment increased NHS-number availability and enabled additional NHS-number-based links, although a small proportion linked to different ONS records following enrichment. NHS number availability and linkage success differed across demographic and social groups, demonstrating differential linkage.

**Conclusions:** A hierarchical deterministic approach combined with targeted validation achieved high linkage accuracy while identifying and addressing differential linkage. This provides a transparent framework for future NTBS–ONS linkage and may be adaptable to other surveillance systems where comparable identifiers are available.

**Key messages:**

- Hierarchical deterministic linkage achieved high precision (>99%) and recall (>96%) when linking tuberculosis notifications to mortality records.
- Validation demonstrated that high linkage accuracy can be achieved using routinely collected identifiers within national surveillance data.
- The validated linkage provides a robust framework for mortality ascertainment and long-term outcome monitoring within tuberculosis surveillance.

## Background

Linkage between infectious disease surveillance systems and national mortality records is fundamental to quantifying long-term mortality, estimating disease burden and evaluating population health outcomes. Tuberculosis (TB) exemplifies this need. An estimated 155 million people worldwide were living after successful treatment for TB in 2020, representing almost 2% of the global population (1). There is increasing recognition that the consequences of TB extend well beyond treatment completion (2). Compared with the general population, TB survivors experience approximately threefold higher mortality (3) together with an increased risk of chronic respiratory disease, cardiovascular disease and disability (4–7). Consequently, post-TB morbidity and mortality are estimated to account for almost half of the total global TB disease burden, measured in disability-adjusted life years (DALYs) (8).

In the UK, TB is a notifiable disease, with suspected and confirmed cases reported through national TB surveillance systems(9). Whilst NTBS provides detailed information on diagnosis, treatment and patient characteristics, surveillance follow-up ends at treatment completion, precluding ascertainment of long-term mortality. Linkage with mortality registries has therefore been fundamental to quantifying long-term mortality following TB treatment internationally (10–13). Similar linkage approaches underpin routine survival estimation in UK cancer registries and HIV surveillance (14,15).

Record linkage between routinely collected health datasets presents methodological challenges, particularly when personal identifiers are incomplete, inaccurate or inconsistently recorded (16). Deterministic linkage applies predefined rules requiring specified identifiers to agree, whereas probabilistic linkage uses the probability of agreement and disagreement across identifiers to estimate whether records belong to the same individual (17). Deterministic approaches are transparent and computationally efficient, but their accuracy depends on careful specification and validation of matching rules (18–20). Probabilistic approaches can accommodate missing or discordant identifiers but may require greater technical infrastructure.

Both approaches have previously been applied to TB surveillance data. Probabilistic linkage has been developed and validated for linking TB notifications with laboratory surveillance data (21), while previous linkage of NTBS data with ONS mortality records has primarily supported ascertainment of TB deaths and estimation of short-term case fatality (22,23). However, a validated framework for deterministic linkage of NTBS with national mortality records to support long-term follow-up has not been reported.

We therefore developed and validated a deterministic framework for linking NTBS records with ONS death registrations. Our objective was to develop a transparent, high-precision and computationally efficient approach using routinely collected identifiers that could support repeat linkage of NTBS to mortality records and be adapted to other surveillance systems with comparable identifiers.

## Methods

### Data sources

TB notifications reported in England between January 2001 and December 2024 were obtained from the 2025 NTBS analytical dataset, produced as described in the published methods of the *National Tuberculosis in England 2025* report (24). The dataset was structured at notification level, with each notification representing a separate episode of TB. An existing NTBS person record identifier enabled multiple notifications belonging to the same individual to be identified.

The dataset includes enrichment from the NHS Personal Demographics Service (PDS) for England and Wales (23). The most recent enrichment was conducted on 22 February 2025 via the Demographic Batch Service (DBS) (24). This provided updated NHS numbers, dates of birth, dates of death, and postcode information where high-quality matches were identified.

Mortality data were obtained from the Office for National Statistics (ONS) mortality database which contains information from death certificates registered in England and Wales (27). This dataset is updated weekly and the extract used for this study was current as of 9 March 2026.

An overview of the linkage, validation and refinement process is presented in Figure 1.

**Figure 1.**
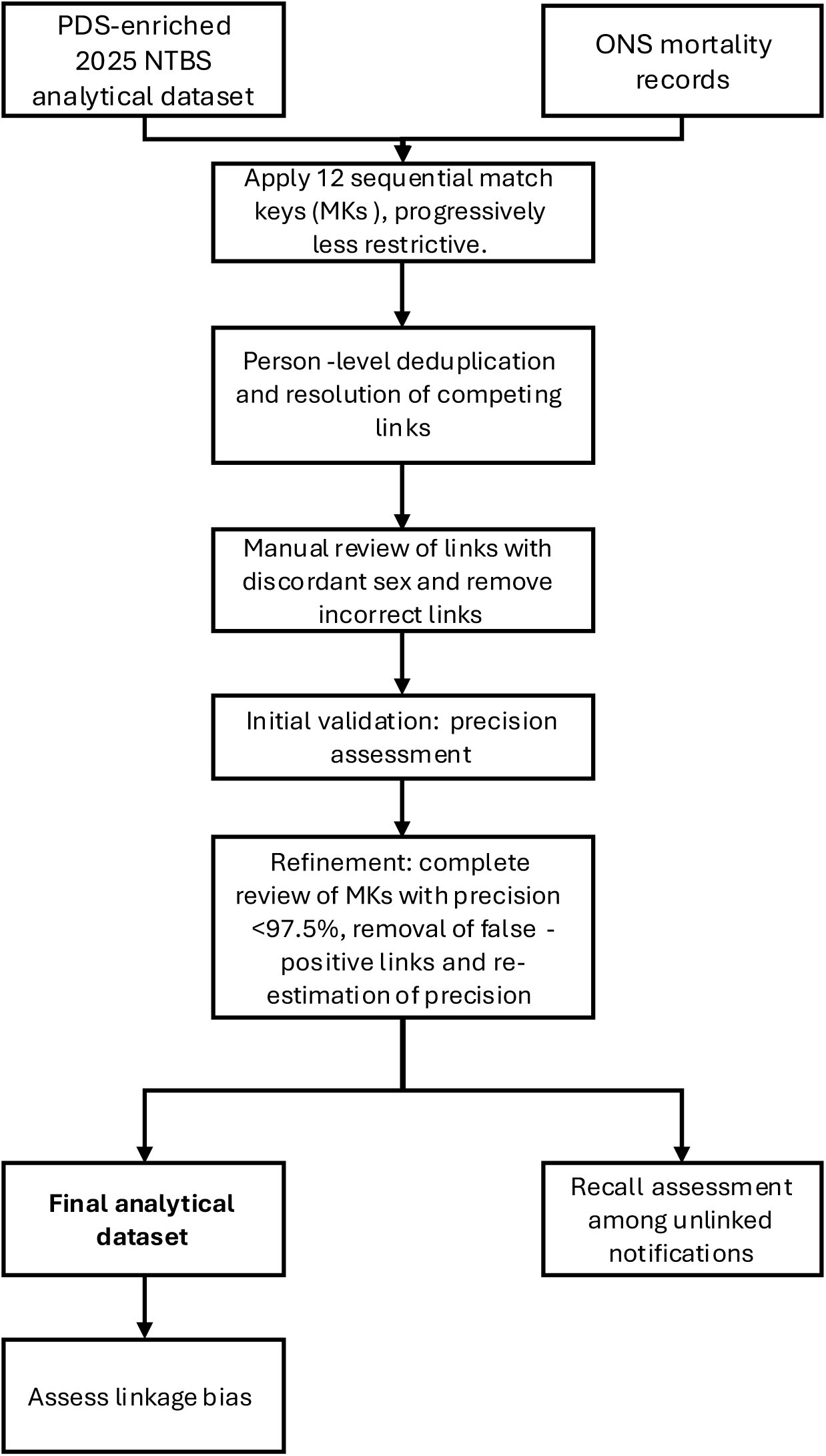
Flow diagram showing deterministic linkage, validation and refinement of the analytical dataset.

### Deterministic linkage

A hierarchical deterministic linkage approach was implemented using 12 match keys (MKs) developed for this study. An MK was defined as a predetermined combination of identifiers that had to agree for a TB notification and ONS mortality record to be considered a match. The 12 MKs were specified a priori by adapting established DBS linkage methods for records containing NHS numbers (28) and ONS Census–PDS linkage methods for records without NHS numbers (29) to identifiers available in both datasets. Additional relaxation beyond MK12 was considered insufficiently specific and not pursued. The MKs were specified before assessment of linkage performance.

MKs were applied sequentially, using progressively less restrictive matching criteria. Early MKs (MK1–4) required concordant NHS numbers, whereas later MKs (MK5–12) relied on combinations of demographic identifiers where NHS numbers were unavailable (**Table 1**).

**Table 1.**
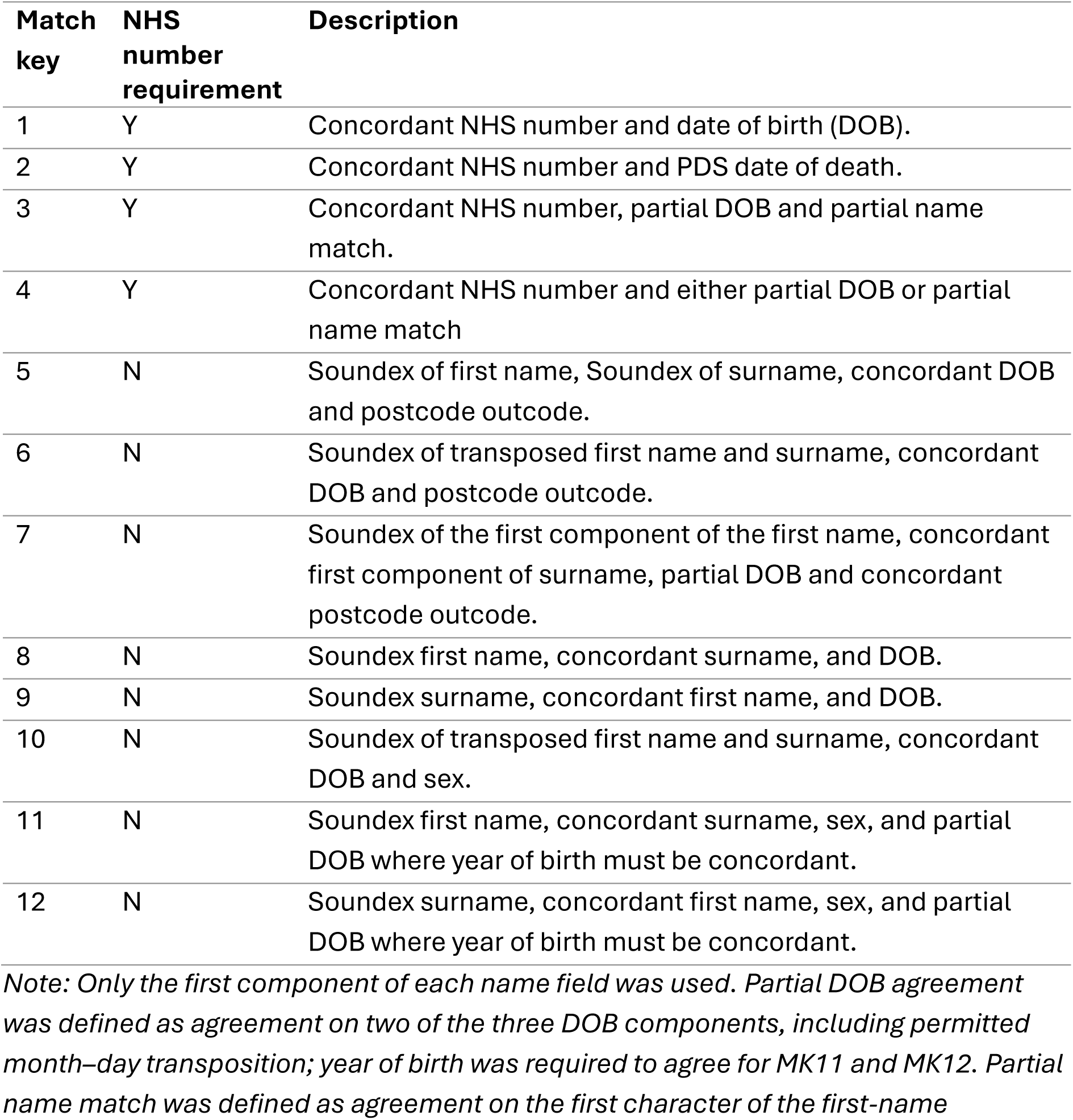
Sequential hierarchical deterministic match keys used for NTBS–ONS record linkage.

| **Match key** | **NHS number requirement** | **Description** |
|---|---|---|
| 1 | Y | Concordant NHS number and date of birth (DOB). |
| 2 | Y | Concordant NHS number and PDS date of death. |
| 3 | Y | Concordant NHS number, partial DOB and partial name match. |
| 4 | Y | Concordant NHS number and either partial DOB or partial name match |
| 5 | N | Soundex of first name, Soundex of surname, concordant DOB and postcode outcode. |
| 6 | N | Soundex of transposed first name and surname, concordant DOB and postcode outcode. |
| 7 | N | Soundex of the first component of the first name, concordant first component of surname, partial DOB and concordant postcode outcode. |
| 8 | N | Soundex first name, concordant surname, and DOB. |
| 9 | N | Soundex surname, concordant first name, and DOB. |
| 10 | N | Soundex of transposed first name and surname, concordant DOB and sex. |
| 11 | N | Soundex first name, concordant surname, sex, and partial DOB where year of birth must be concordant. |
| 12 | N | Soundex surname, concordant first name, sex, and partial DOB where year of birth must be concordant. |

*Note: Only the first component of each name field was used. Partial DOB agreement was defined as agreement on two of the three DOB components, including permitted month–day transposition; year of birth was required to agree for MK11 and MK12. Partial name match was defined as agreement on the first character of the first-name* [illegible]

Sex was not included in MK1–7 to maximise sensitivity. Records with discordant sex were instead manually reviewed after deduplication, and links considered incorrect were removed.

Linkage was conducted within the UK Health Security Agency Enterprise Data and Analytics Platform (EDAP), using Amazon Redshift, with linkage algorithms implemented in SQL.

### Deduplication and review of competing links

Linkage outputs were assessed at NTBS person level to identify cases where one NTBS person linked to multiple ONS records (one-to-many), multiple NTBS person records linked to the same ONS record (many-to-one), or multiple NTBS person records linked to multiple ONS records (many-to-many). These links were manually reviewed using additional identifiers and incorrect links were removed, ensuring all retained NTBS person–ONS record links were one-to-one (**Supplementary Methods 1)**.

### Validation of linkage

#### Precision

Following person-level resolution and review of links with discordant sex, a random sample of 220 linked NTBS notification–ONS record pairs within each MK underwent clerical review (manual checking). Where an MK contained fewer than 220 links, all links were reviewed. Additional identifiers were reviewed and links were classified as true positives (correct links) or false positives (incorrect links).

Precision was calculated as:

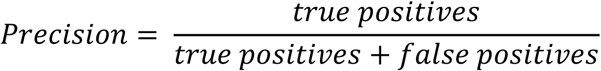

The precision calculated from each MK sample was applied to the total number of links assigned to that MK to estimate the number of true-positive links. Estimated true-positive links were then summed across all MKs to calculate overall weighted precision. Precision estimates were reported with 95% confidence intervals calculated using the Agresti–Coull method (30). Full sampling and calculation procedures are provided in **Supplementary Methods 2**.

##### Post review refinement of the analytical dataset: removal of false positives

After completing the precision assessment, all links within MKs with precision below a prespecified threshold of 97.5% underwent complete clerical review. This threshold was selected to prioritise review of likely false-positive links. Confirmed false-positive links were removed from the analytical dataset and overall precision was re-estimated (**Supplementary Methods 3)**.

#### Recall

Recall (or sensitivity) was assessed among notifications remaining unlinked after removal of false positives from the precision assessment. Relaxed matching criteria were used to identify plausible missed links (**Table 2**).

**Table 2.** Recall search criteria used to identify potential missed links.

| Stratum | Candidate search rule |
|---|---|
| **A** | Concordant NHS number only. |
| **B** | Concordant DOB, postcode outcode and Soundex agreement on either first name or surname; discordant NHS numbers allowed. |
| **C** | Concordant DOB, and either exact agreement on first name or surname; discordant NHS numbers allowed. |

A sample of distinct notifications identified within each stratum underwent clerical review to determine whether they represented false-negative links or true non-links. Confirmed false-negative links identified during recall assessment were used to estimate recall but were not incorporated into the final linked dataset. The proportion of missed links identified in each reviewed sample was applied to all eligible notifications within the corresponding stratum to estimate the total number of missed links. These estimates were combined across the recall strata to calculate overall recall:

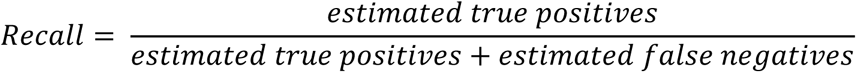

Full search criteria, sampling procedures and calculations are provided in

**Supplementary Methods 4**.

### Assessment of linkage bias

Because the linkage relied predominantly on NHS numbers, potential linkage bias was assessed by examining whether NHS-number availability and linkage success differed systematically across population groups. Trends in NHS-number availability and the proportion of linked notifications identified through NHS-number-based MKs were examined by TB notification year. NHS-number availability was also compared by age, sex, ethnicity, UK-born status and recorded social risk factors, including homelessness, imprisonment, alcohol misuse, drug misuse and asylum-seeker status; missing values were retained as a separate category. Finally, linkage success was compared between notifications with and without an NHS number using a chi-squared test.

### Sensitivity analysis of PDS enrichment

A sensitivity analysis assessed the impact of prior PDS enrichment on NTBS–ONS linkage by comparing linkage outcomes using original NTBS and PDS-enriched identifiers among notifications where PDS supplied a previously missing NHS number or where NHS numbers were discordant (**Supplementary Methods 5**).

### Comparison of death records across NTBS, PDS and ONS

Agreement in recorded death status was compared across NTBS, PDS and ONS. The numbers and overlap of deaths recorded in each source were summarised descriptively.

## Results

### Linkage outcomes

A total of 151,965 TB notifications were reported in England between 2001 and 2024, of which 28,060 initial notification-level links to ONS mortality records were identified (**Table 3**). Following person-level resolution of duplicate and competing links, review of links with discordant sex, and removal of confirmed false-positive links, 26,937 ONS-linked notifications were retained (17.7% of all notifications) (**Supplementary Table S2)**.

**Table 3.** Summary of NTBS-ONS linkage outcomes following deterministic linkage and clerical review.

| **Match key** | **Initial number of links** | **Final number of links** | **% of total number of retained links (n=26,937)** |
|---|---|---|---|
| **1** | 24,079 | 24,073 | 89.37 |
| **2** | 475 | 474 | 1.76 |
| **3** | 18 | 18 | 0.07 |
| **4** | 51 | 49 | 0.18 |
| **5** | 1,302 | 1,288 | 4.78 |
| **6** | 112 | 112 | 0.42 |
| **7** | 319 | 317 | 1.18 |
| **8** | 642 | 365 | 1.36 |
| **9** | 127 | 116 | 0.43 |
| **10** | 80 | 39 | 0.14 |
| **11** | 649 | 79 | 0.29 |
| **12** | 206 | 7 | 0.03 |
| **Total** | 28,060 | 26,937 | 100 |

*Note: Following initial linkage, duplicate and competing links were resolved at the person level, links with discordant sex were reviewed, and confirmed false-positive links were removed to derive the final number of retained links. The number of links retained at each stage is detailed in* ***Supplementary Table S2****.*

### Deduplication and resolution of competing links

The 28,060 initial NTBS notification–ONS record links corresponded to 27,403 distinct NTBS person–ONS record combinations. Following person-level resolution of duplicate and competing links, 420 NTBS person–ONS record combinations were removed, leaving 26,983 one-to-one NTBS person–ONS record links, representing 27,356 TB notifications (**Supplementary Table S3**).

Subsequently, 56 notification-level links with discordant recorded sex were manually reviewed; four were removed as incorrect, leaving 27,352 notification-level links (**Supplementary Table S2**).

### Validation

#### Precision

A total of 1,789 records were sampled for clerical review across all MKs. Of these, 313 were identified as false positives, primarily arising from MK 8–12, which employ less restrictive matching criteria (**Table 4**).

**Table 4.**
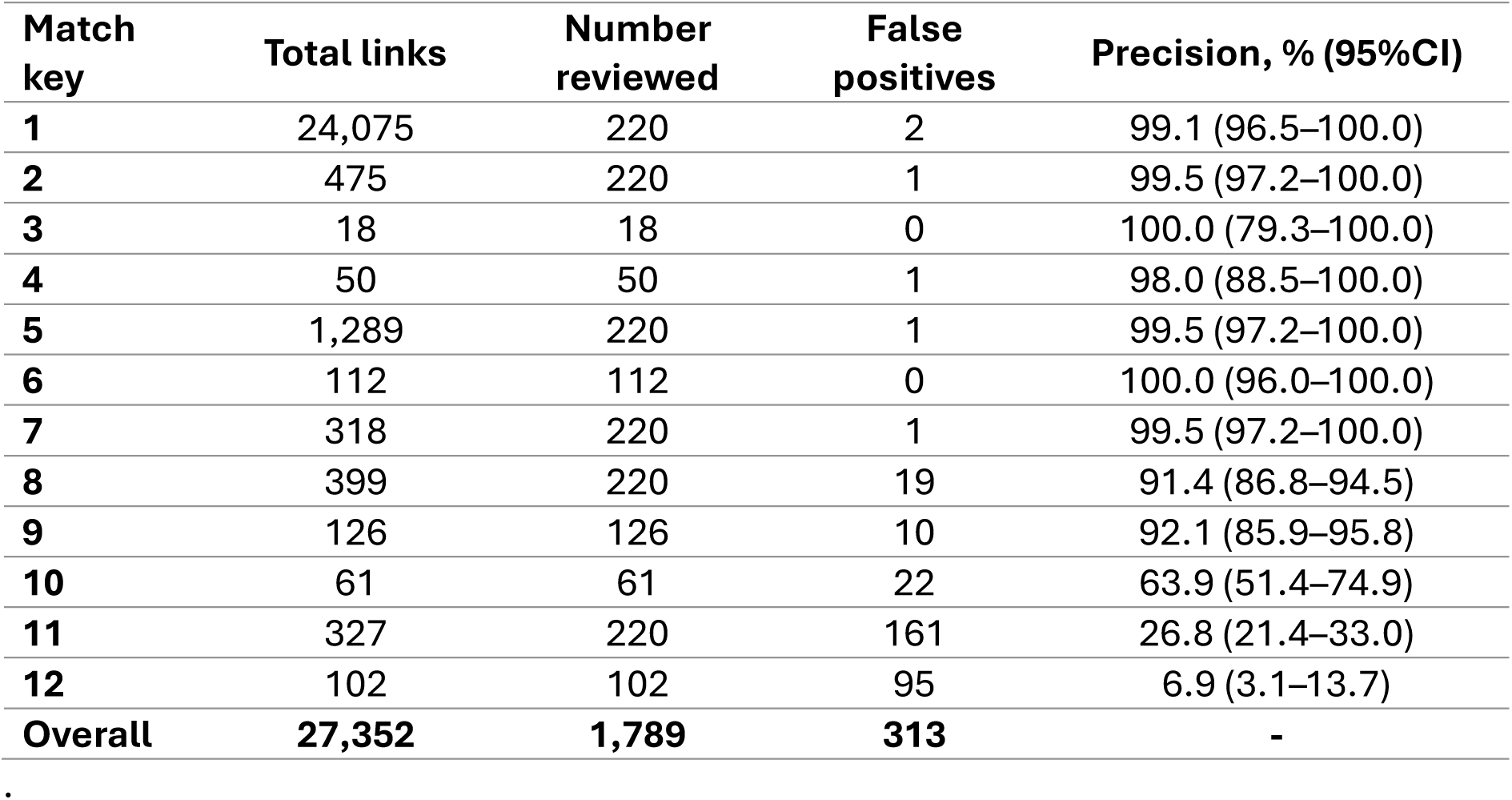
Initial precision assessment of deterministic match keys.

Weighted by the number of links contributed by each MK, overall linkage precision was estimated at 97.70% (95% CI 95.02%-98.69%) (**Supplementary Table S4)**.

##### Post precision refinement of the analytical dataset

Initial precision assessment identified that MK 8–12 had estimated precision below the prespecified threshold of 97.5%. The remaining 286 unsampled links within these MKs therefore underwent complete clerical review. This review identified a further 102 false-positive links, increasing the total number of confirmed false-positive links from 313 to 415, all of which were removed from the analytical dataset. As a result, the estimated weighted precision of the analytical dataset increased from 97.7% to 99.2% (95% CI 96.8%–100.0%) (**Supplementary Table S5**).

#### Recall

Recall assessment included 13,107 eligible notifications across three strata. Of 458 notifications reviewed, 133 contained a confirmed false-negative link. Applying the sampling-group-specific false-negative proportions to the corresponding eligible populations produced an estimated 527 missed links and an overall recall of 98.1% (95% CI 95.4–98.6%) (**Supplementary Table S6**).

### Linkage bias

NHS number completeness increased substantially over the study period, from 64.9% in 2001 to 98.9% in 2024 (**Figure 2A**). Correspondingly, the proportion of linked notifications identified using NHS-number-based MKs increased from 81.7% to 99.2% (**Figure 2B**). Overall, more than 90% of all linked notifications were matched using NHS-number-based MKs (MK1–4).

**Figure 2.**
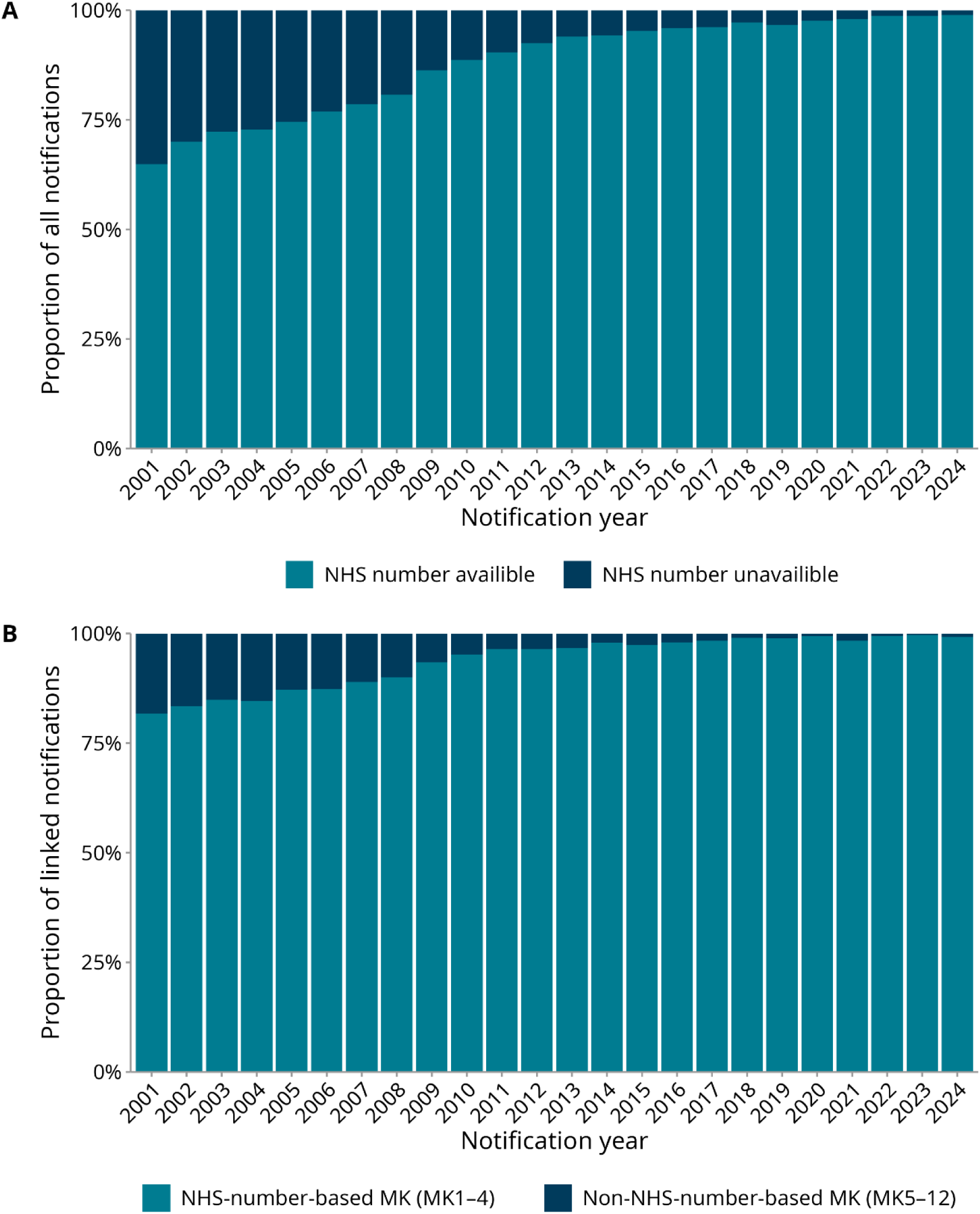
NHS number availability (A) and use of NHS-number-based match keys (B) by notification year, England, 2001–2024. Note: (A) Proportion of TB notifications with an NHS number available, by notification year. (B) Proportion of linked TB notifications identified using NHS-number-based (MK1– 4) and non-NHS-number-based (MK5–12) match keys, by notification year (**Supplementary Table S7**).

NHS number availability varied by demographic and social characteristics (**Supplementary Table S8**), suggesting that the potential for linkage bias differed across population subgroups. The proportion of notifications without an NHS number was higher among non-UK-born than UK-born individuals (14.8% vs 7.6%), and among those with a recorded history of homelessness (12.8% vs 5.5%) or imprisonment (10.5% vs 5.7%) compared with those without. NHS number absence was also more common among notifications with missing social risk factor information.

Consistent with this, notifications containing an NHS number were significantly more likely to be successfully linked than notifications without an NHS number (18.7% vs 11.5%; χ² p” 0.001; **Table 5**).

**Table 5.** Linkage success according to NHS number availability.

| **NHS number availability** | **Linked**<br>**n (%)** | **Not linked**<br>**n (%)** | **p-value** |
|---|---|---|---|
| **NHS number present** | 24,617 (18.7) | 107,059 (81.3) | <0.001 |
| **NHS number absent** | 2,325 (11.5) | 17,964 (88.5) |  |

*Note: p-value was calculated using Pearson’s chi-squared test.*

### Sensitivity analysis of PDS enrichment

Among 35,072 notifications for which PDS enrichment added an NHS number, 441 (1.3%) linked only after enrichment. Conversely, 1,158 (3.3%) linked only using pre-enrichment identifiers, however 1,046 (90.3%) of these links arose through the lower-precision MK11–MK12. A further 104 notifications (0.3%) linked to different ONS records before and after enrichment.

Among 685 notifications with discordant original and PDS-derived NHS numbers, 149 (21.8%) linked only using the PDS-derived number and five (0.7%) linked to different ONS records. No notifications linked only using the original NHS number (**Supplementary Result 5, Supplementary Table SG)**.

### Comparison of death records across NTBS, PDS, and ONS

Comparison of death status across NTBS, PDS and ONS showed substantial but incomplete agreement between sources. Differences appeared to reflect variation in underlying source systems rather than linkage failure (**Supplementary Result 6**).

## Discussion

### Principal findings

This study developed and validated, to our knowledge, the first published national deterministic linkage framework linking NTBS notifications to ONS mortality records in England. Validation indicated very high estimated precision and recall, while supplementary deterministic MKs enabled valid death records to be identified among individuals without NHS numbers. Assessment of linkage bias demonstrated that NHS number availability varied over time and across demographic and social groups, highlighting the potential for differential linkage. These findings demonstrate the value of combining formal linkage validation with assessment of identifier completeness and differential linkage when developing surveillance-based linkage frameworks (27).

### Performance and validation of the linkage framework

The combination of high NHS number completeness, PDS identifier enrichment and hierarchical deterministic matching contributed to the high linkage performance achieved.

The linkage framework is consistent with recent ONS deterministic linkage evaluations, which similarly employed hierarchical MKs, clerical review and formal assessment of linkage quality and bias (29,30). The high accuracy achieved in the present study is consistent with findings from these evaluations and broader evidence that deterministic linkage performs well when reliable identifiers are available.

A key strength of deterministic linkage is that it provides a transparent and reproducible framework, with explicit matching rules that can be consistently applied over time, making it well suited to routine national surveillance and repeat analyses. Validation was integrated into the linkage refinement process, with precision estimates guiding targeted clerical review to remove false-positive matches while retaining valid links. This refinement process was feasible because the number of links requiring review was relatively small compared with large population-scale linkage studies undertaken by ONS.

### Differential linkage bias and implications for mortality ascertainment

Although linkage quality was high overall, evidence of differential linkage remained. NHS number absence was more common among non-UK-born individuals and those with a recorded history of homelessness or imprisonment. Differences in identifier availability, alongside substantial variation in missing social risk factor information, suggest that the potential for linkage bias may vary across population subgroups. Similar patterns have been reported elsewhere, highlighting the potential for linkage processes to affect representativeness (27,29). These findings emphasise that evaluation of linkage quality should extend beyond traditional measures of precision and recall to include assessment of linkage bias.

Lower-precision MKs were retained because, despite generating more false-positive links, they identified genuine matches among individuals lacking NHS numbers. Excluding them could have exacerbated differential linkage and the potential for selection bias. Instead, targeted clerical review removed false-positive links while retaining valid matches, increasing the estimated precision of the analytical dataset.

Despite the high linkage accuracy, some mortality under-ascertainment may remain because linkage performance depends on the completeness of the underlying mortality data. Deaths occurring outside England and Wales that are not subsequently notified to the NHS may be absent from both ONS and PDS records and therefore cannot be identified through linkage. This source of under-ascertainment should be distinguished from linkage error when interpreting estimates derived from linked surveillance data.

### Strengths and limitations

Strengths include the transparent description of the linkage methodology, a large national surveillance dataset spanning 24 years, and formal assessment of precision, recall and linkage bias.

PDS enrichment improved identifier completeness and enabled additional links through high-precision NHS-number-based MKs, while sensitivity analyses demonstrated that changes to the identity of linked ONS records were uncommon. Nevertheless, the identification of one erroneous linkage resulting from incorrect PDS enrichment highlights the need for caution and supports consideration of both original and enriched identifiers in future linkage exercises. Investigation of discordant mortality records indicated that discrepancies between PDS and ONS largely reflected differences in the underlying source systems rather than linkage errors. These findings suggest that PDS and ONS should be regarded as complementary rather than interchangeable sources of mortality information.

Several limitations should also be acknowledged. First, the linkage relied exclusively on deterministic methods. Although deterministic linkage offers transparency, reproducibility and ease of implementation within routine surveillance systems, probabilistic linkage may recover additional true matches where identifiers are incomplete (or erroneous), implementation was constrained by technical, computational and information-governance requirements within the EDAP environment. Large-scale population linkage programmes, including the Census 2021 linkage programme, have demonstrated the value of probabilistic methods in settings where linkage relies primarily on combinations of names, dates of birth and addresses rather than unique identifiers(31). However, given the high availability of NHS numbers, the use of PDS-enriched identifiers, and the high precision and recall achieved, the incremental benefit of probabilistic linkage in the present study was likely to be relatively small, although this could not be directly assessed.

Second, the linkage was constrained by the identifiers available within the source datasets and the deterministic matching rules applied. The recall assessment identified a small number of missed matches resulting from the exclusion of middle names and secondary NHS numbers from the MKs. NHS-number-based MKs required exact agreement, and discordant NHS numbers were not explicitly accommodated within the matching rules. However, the recall assessment also identified plausible missed links where NHS numbers differed but name, date of birth and postcode outcode were concordant, indicating that allowing such combinations, with appropriate validation, may improve recall in future linkage iterations. Middle names were considered during clerical review, and the high overall recall suggests that the impact of these limitations was relatively small.

Third, clerical review inevitably involves an element of subjective judgement. Reviews were conducted primarily by a single reviewer with uncertain cases discussed with a second reviewer, to improve consistency. Nevertheless, the conservative review strategy may also have resulted in a small number of genuine matches being excluded where insufficient evidence existed to confirm linkage with confidence.

## Conclusion

This study demonstrates that high-accuracy deterministic linkage between national TB surveillance and mortality records can be achieved despite incomplete identifiers. Combining PDS-enriched identifiers, hierarchical match keys and targeted clerical review enabled high precision and recall while retaining valid links identified through lower-precision match keys. Importantly, the findings highlight the value of assessing differential linkage alongside traditional measures of linkage quality. The resulting reproducible framework supports future NTBS-ONS linkage and, although the specific match keys were developed for the identifiers available within NTBS and ONS mortality records, may be adapted for linkage, validation and assessment of linkage bias in other surveillance and administrative data settings.

## Supporting information

Supplementary Methods and Results

## Ethics approval

The study for which this data linkage was conducted was approved by the Health, Biological and Psychological Sciences Research Ethics Committee at University of Leicester (Project ID 1760) and received UKHSA governance approval (RITM0158729) (32).

Individual consent was not required. Data linkage was undertaken within UKHSA using routinely collected public health surveillance data before provision of the study dataset. The final analytical dataset was pseudonymised and contained no personal identifiable information.

## Acknowledgements

The authors thank members of the ONS Data Linkage Expert Group for constructive feedback on the linkage methodology and manuscript, including discussion at an expert group meeting and comments on a draft version of the manuscript.

## Author contributions

OP designed methodology, conducted data curation, analysis and wrote the original draft, AO contributed to conceptualisation, methodology and writing the original draft, DZ and JM contributed to supervision, and writing – review and editing, MP contributed to conceptualisation, supervision, funding acquisition, and writing – review and editing, SC contributed to conceptualisation, methodology, supervision, and writing – review and editing. All authors reviewed and approved the final manuscript.

## Supplementary data

Supplementary data are available at *IJE* online.

## Data availability

The data underpinning this study is held by the UKHSA. Access to the data may be considered through UKHSA’s established data access governance processes and is subject to the necessary approvals. Further information on the data access process is available at: https://www.gov.uk/government/publications/accessing-ukhsa-protected-data/accessing-ukhsa-protected-data.

## Use of Artificial intelligence (AI) tools

AI-assisted tools were used to support code development and manuscript editing, including improvements to clarity and presentation. All content was reviewed and verified by the authors, who take full responsibility for the final manuscript.

