## Supplementary Methods and Results for "A deterministic framework for linkage of infectious disease surveillance and national mortality records: development and validation using tuberculosis surveillance data"

This supplement provides detailed linkage specifications, adjudication procedures, validation methods and sensitivity analyses supporting the main manuscript. The primary analysis linked National Tuberculosis Surveillance System notifications in England from 2001 to 2024 to Office for National Statistics mortality records available up to 9 March 2026.

### Abbreviations

CI, confidence interval; DOB, date of birth; DBS, Demographic Batch Service; EDAP, Enterprise Data and Analytics Platform; FN, false negative; FP, false positive; MK, match key; NHS, National Health Service; NTBS, National Tuberculosis Surveillance System; ONS, Office for National Statistics; PDS, Personal Demographics Service; TN, true negative; TP, true positive.

### Supplementary Methods

#### Supplementary Methods 1. Deduplication and review of competing links

Initial NTBS notification-level links were grouped by NTBS-person record and ONS-record and classified into four structures: one NTBS person linked to one ONS record; one NTBS person linked to multiple ONS records; multiple NTBS person records linked to one ONS record; and multiple NTBS person records linked to multiple ONS records.

Duplicate and competing links were manually reviewed using additional identifiers, including extended name fields, full date of birth, sex, postcode fields and country of birth. The objective was to retain no more than one ONS mortality record for each NTBS person. Links were removed when the available identifiers suggested that the records belonged to different individuals or did not provide sufficient evidence to confirm a match.

Because linkage was conducted at notification level, identifiers could vary between notifications belonging to the same person. These values were not harmonised before linkage, allowing alternative identifiers to contribute to matching and reducing the risk of missed links. Consequently, different notifications could link to the same or different ONS records and could satisfy different MKs. Where notifications belonging to the same NTBS person linked to different ONS records, the MK through which each link was identified, and the additional available identifiers were considered to determine the most plausible match. If a single ONS record could not be determined, the competing links were removed.

Where multiple notifications belonging to the same NTBS person linked to the same ONS record through different MKs, the most specific MK identified across those notifications was assigned as a derived person-level MK. For example, if two notifications belonging to the same person linked to the same ONS record through MK1 and MK5, MK1 was assigned as the person-level MK for both notifications. The original notification-level MK was retained separately and used for the validation analyses. The derived person-level MK was created only to allow users of the final analytical dataset to identify the most specific linkage evidence available for each person and was not used for any validation assessment.

#### Supplementary Methods 2. Precision

Following deduplication and review of links with discordant sex, a random sample of links within each MK underwent clerical review (manual checking). Sample sizes were calculated using a standard method for estimating a proportion, consistent with ONS linkage methodology (27,28). The calculation assumed an expected precision of 95%, a 95% confidence level and an absolute margin of error of approximately ±3 percentage points. This produced a target sample of 220 links per MK. Where fewer than 220 links were available, all links were reviewed.

Clerical review compared extended name fields, date of birth, postcode, sex and country of birth were compared, and links were classified as true positives (correct links) or false positives (incorrect links). Precision of each match key was calculated as:

$$Precision = \frac{true\ positives}{true\ positives + false\ positives}$$

The precision calculation for each MK was applied to all deduplicate links assigned to that MK. The estimated number of true positives within each match-key was calculated as:

$$estimated\ true\ positives = precision\ \times\ links$$

Lastly, the estimated number of true positives were summed across all match keys and used to calculate the overall weighted precision as:

$$overall\ weighted\ precision = \frac{sum\ of\ estimated\ true\ positives}{sum\ of\ all\ links}$$

Estimated precision was accompanied by 95% confidence intervals (CIs) calculated using the Agresti-Coull method. The lower and upper confidence limits for each MK were applied to the total number of links assigned to that MK to estimate the corresponding lower and upper numbers of true-positive links. These estimates were summed across all MKs and divided by the total number of links to derive the overall weighted 95% confidence interval (29).

#### Supplementary Methods 3. Post review refinement of the analytical dataset: removal of false positives

After completing the precision assessment, all links within match keys with estimated precision below 97.5% underwent complete manual review. This was an operational threshold to identify match keys requiring further review while avoiding unnecessary review of match keys already demonstrating high estimated precision. Any confirmed false-positive links identified during this process were removed from the analytical dataset.

Following refinement, weighted precision was re-estimated. MK3, MK4 and MK6 had already undergone complete review because they contained fewer than 220 links. Following the additional review, MK8–MK12 had also been completely reviewed. All retained links in these MKs were therefore treated as confirmed true positives. For partially reviewed MKs, reviewed links retained as true positives were combined with the estimated number of true positives among unreviewed links, calculated using the corresponding MK-specific precision estimate. Overall refined precision was calculated by dividing the sum of confirmed and estimated true-positive links across all MKs by the total number of retained links.

MK8–MK12 were retained within the linkage strategy because they provided opportunities to identify matches among records without NHS numbers. Completely reviewing these MKs allowed valid links to be retained while removing confirmed false positives, thereby supporting linkage completeness and reducing potential bias associated with variation in NHS number availability.

### Supplementary Methods 4. Recall

Recall (or sensitivity) was assessed among notifications remaining unlinked after deduplication, person-level resolution and removal of false positives following precision. The assessment was designed to estimate the number of missed links to validate the linkage method using relaxed deterministic searches (Supplementary Table S1).

**Supplementary Table S1 - Recall search criteria**

| Stratum | Candidate search rule | Detail strata | Purpose |
|---|---|---|---|
| **A** | Concordant NHS number only. | | Assesses missed links despite a shared unique identifier. |
| **B** | Concordant DOB, postcode outcode and either Soundex of first name or Soundex of surname; discordant NHS numbers allowed. | B1: Soundex first name and surname<br>B2: Soundex first name only<br>B3: Soundex surname only | Identifies potential matches where DOB and postcode agree but one or both names contain spelling variations or different name components, including nicknames, middle names or maiden names and/or NHS number disagreed or changed. |
| **C** | Concordant DOB, and either concordant first name or concordant surname; discordant NHS numbers allowed. | C1: Exact first name and surname<br>C2: Exact first name only<br>C3: Exact surname only | Identifies potential matches where DOB and one or both names agree but postcode may have changed and/or NHS number disagreed or changed. |

A target sample of 220 distinct notifications was selected within each main stratum, consistent with the sampling approach used for the precision assessment. Where a main stratum contained fewer than 220 eligible notifications, all notifications were reviewed. Within strata B and C, the target sample was allocated proportionally across the constituent detail strata and notifications were randomly sampled. Where a sampled notification produced multiple candidate ONS records, all candidates were reviewed.

Clerical review compared extended name fields, date of birth, postcode, sex and country of birth. A sampled notification was classified as containing a false-negative link when one candidate ONS record was judged to represent the same individual but had not been identified by the primary linkage. Notifications for which none of the candidate ONS records represented the same individual were classified as true negatives.

Within each detail stratum, the false negative proportion was calculated as:

$$false\ negative\ proportion = \frac{false\ negatives}{false\ negatives + true\ negatives}$$

The estimated number of false-negative links in each detail stratum was calculated as:

$$Estimated\ false\ negatives = false\ negative\ proportion\ \times eligible\ notifications$$

Estimated false negatives were summed across all detail strata. Overall recall was calculated using the estimated number of true-positive links from the precision assessment and the estimated number of false-negative links:

$$Recall = \frac{estimated\ true\ positives}{estimated\ true\ positives + estimated\ false\ negatives}$$

False-negative estimates and 95% CIs were calculated and weighted across sampling groups using the same approach described for the precision assessment.

#### Supplementary Methods 5. Sensitivity analysis of PDS enrichment

PDS enrichment was used before NTBS-ONS linkage to improve the completeness and quality of NHS number information. Sensitivity analyses assessed notifications for which enrichment either added or changed the NHS number available for linkage.

First, 35,072 notifications had no NHS number recorded before PDS enrichment and subsequently obtained a PDS-derived NHS number. Linkage was repeated using the pre-enrichment data, in which NHS number was missing, and compared with the primary linkage using the PDS-derived NHS number. Second, 685 notifications had an original NTBS NHS number that differed from the PDS-derived NHS number. For those records, the primary linkage used the PDS-derived number and the sensitivity analysis repeated linkage using the original pre-PDS number.

The same deterministic linkage algorithm was applied in each comparison. The ONS mortality data were restricted to records available before 9 March 2026 so that both runs used the same candidate pool. Outcomes were classified as unlinked under both approaches, linked to the same ONS record, linked under only one approach, or linked to different ONS records. Match-key distributions were examined for discordant outcomes.

### Supplementary Results

#### Supplementary Result 1. Deduplication and review of competing links

**Supplementary Table S2 - Number of notification-level links retained at each stage of the linkage and refinement process**

| MK | Initial notification-level links | Links after person-level resolution and deduplication | Links with discordant sex reviewed | Links after discordant-sex review | Final number of links after removal of false positives |
|---|---|---|---|---|---|
| **1** | 24,079 | 24,078 | 24 | 24,075 | 24,073 |
| **2** | 475 | 475 | 2 | 475 | 474 |
| **3** | 18 | 18 | 1 | 18 | 18 |
| **4** | 51 | 50 | 0 | 50 | 49 |
| **5** | 1,302 | 1,289 | 0 | 1,289 | 1,288 |
| **6** | 112 | 112 | 2 | 112 | 112 |
| **7** | 319 | 318 | 2 | 318 | 317 |
| **8** | 642 | 400 | 22 | 399 | 365 |
| **9** | 127 | 126 | 3 | 126 | 116 |
| **10** | 80 | 61 | NA | 61 | 39 |
| **11** | 649 | 327 | NA | 327 | 79 |
| **12** | 206 | 102 | NA | 102 | 7 |
| **Total** | **28,060** | **27,356** | **56** | **27,352** | **26,937** |

*Note: Counts are notification-level links. After the removal of false positives, a person-level MK was created resulting in the reallocation of MK for 63 notifications but this did not change the total number of links (**Supplementary methods 1**).*

Supplementary Table S3 - Link structures before and after deduplication and resolution of competing links

| Link structure | Before review n | Before review % | After resolution n | After resolution % |
|---|---|---|---|---|
| **One NTBS person to one ONS record** | 26,576 | 96.98 | 26,983 | 100.00 |
| **One NTBS person to multiple ONS records** | 57 | 0.21 | 0 | 0.00 |
| **Multiple NTBS person records to one ONS record** | 732 | 2.67 | 0 | 0.00 |
| **Multiple NTBS person records to multiple ONS records** | 38 | 0.14 | 0 | 0.00 |
| **Total linked NTBS person records** | 27,403 | 100.00 | 26,983 | 100.00 |

*Note: After resolution, retained links from complex clusters contribute to the one-to-one category; duplicate or implausible competing links were removed. The increase in one-to-one links therefore reflects reclassification of resolved clusters, not an increase in total candidate links*

### Supplementary Result 2. Precision

**Supplementary Table S4 - Precision results**

| Match key | Total links | Total links sampled for review | True positives (TP) | False positives (FP) | Precision (TP/TP+FP) | Estimated TP (95% CI) |
|---|---|---|---|---|---|---|
| **1** | 24,075 | 220 | 218 | 2 | 99.1 (96.5–100.0) | 23856.1 (23239.6–24067.0) |
| **2** | 475 | 220 | 219 | 1 | 99.5 (97.2–100.0) | 472.8 (461.7–475.0) |
| **3** | 18 | 18 | 18 | 0 | 100.0 (79.3–100.0) | 18.0 (14.3–18.0) |
| **4** | 50 | 50 | 49 | 1 | 98.0 (88.5–100.0) | 49.0 (44.3–50.0) |
| **5** | 1,289 | 220 | 219 | 1 | 99.5 (97.2–100.0) | 1283.1 (1253.0–1289.0) |
| **6** | 112 | 112 | 112 | 0 | 100.0 (96.0–100.0) | 112.0 (107.5–112.0) |
| **7** | 318 | 220 | 219 | 1 | 99.5 (97.2–100.0) | 316.6 (309.1–318.0) |
| **8** | 399 | 220 | 201 | 19 | 91.4 (86.8–94.5) | 364.5 (346.5–376.9) |
| **9** | 126 | 126 | 116 | 10 | 92.1 (85.9–95.8) | 116.0 (108.2–120.7) |
| **10** | 61 | 61 | 39 | 22 | 63.9 (51.4–74.9) | 39.0 (31.3–45.7) |
| **11** | 327 | 220 | 59 | 161 | 26.8 (21.4–33.0) | 87.7 (69.9–108.1) |
| **12** | 102 | 102 | 7 | 95 | 6.9 (3.1–13.7) | 7.0 (3.2–14.0) |
| **Total** | 27,352 | 1,789 | 1,476 | 313 | | 26721.9 (25988.7–26994.4) |

Following the calculation of precision for each MK, the sum of MK calculated TP was 26721.9 (25988.7–26994.4). Out of a total 27,352 links, this resulted in a weighted precision of 97.70% CI [95.02%-98.69%].

**Supplementary Result 3. Post review refinement of analytical dataset and re-estimation of precision**

**Supplementary Table S5 - Refinement of analytical dataset and re-estimation of precision**

| MK | Initial sample | Additional links for full clerical review | Extra FP | Total FP identified removed from dataset | Final retained links | Review status | Unreviewed links | Estimated TPs after refinement, 95%CI | Re-estimated precision, % (95%CI) |
|---|---|---|---|---|---|---|---|---|---|
| **1** | 220 | - | - | 2 | 24073 | Partial | 23,855 | 23,856.1 (23,245.2–24,065.1) | 99.1 (96.6–100.0) |
| **2** | 220 | - | - | 1 | 474 | Partial | 255 | 472.8 (466.9–474.0) | 99.8 (98.5–100.0) |
| **3** | 18 | - | - | 0 | 18 | Complete | 0 | 18.0 (18.0–18.0) | 100.0 (100.0–100.0) |
| **4** | 50 | - | - | 1 | 49 | Complete | 0 | 49.0 (49.0–49.0) | 100.0 (100.0–100.0) |
| **5** | 220 | - | - | 1 | 1288 | Partial | 1,069 | 1,283.1 (1,258.2–1,288.0) | 99.6 (97.7–100.0) |
| **6** | 112 | - | - | 0 | 112 | Complete | 0 | 112.0 (112.0–112.0) | 100.0 (100.0–100.0) |
| **7** | 220 | - | - | 1 | 317 | Partial | 98 | 316.6 (314.3–317.0) | 99.9 (99.1–100.0) |
| **8** | 220 | 179 | 15 | 34 | 365 | Complete | 0 | 365.0 (365.0–365.0) | 100.0 (100.0–100.0) |
| **9** | 126 | - | - | 10 | 116 | Complete | 0 | 116.0 (116.0–116.0) | 100.0 (100.0–100.0) |
| **10** | 61 | - | - | 22 | 39 | Complete | 0 | 39.0 (39.0–39.0) | 100.0 (100.0–100.0) |
| **11** | 220 | 107 | 87 | 248 | 79 | Complete | 0 | 79.0 (79.0–79.0) | 100.0 (100.0–100.0) |
| **12** | 102 | - | - | 95 | 7 | Complete | 0 | 7.0 (7.0–7.0) | 100.0 (100.0–100.0) |
| **Overall** | **1,789** | **286** | **102** | **415** | **26937** | **—** | **25277** | **26,713.7 (26,069.5–26,929.1)** | **99.2 (96.8–100.0)** |

**Re-estimation of TPs=confirmed reviewed TP+(unreviewed links × initial precision), **Re-estimation of precision= estimated TP/ final retained links*

Note: The initial precision sample included 1,789 links, of which 313 were classified as FPs. FPs were concentrated in MK8-MK12, where the primary rules relied on less restrictive demographic combinations. Weighted overall precision before refinement was 97.70% (95% CI 95.02%-98.71%).

Because estimated precision for MK8-MK12 was below 97.5%, the remaining 286 unsampled links in these strata underwent complete review. A further 102 FPs were identified, giving 415 confirmed FPs removed across the initial and additional reviews. The refined dataset contained 26,937 linked notifications before the recall performance estimate was applied. Re-estimated overall precision was approximately 99.2% (95% CI 96.8–100.0).

### Supplementary Result 4. Recall

**Supplementary Table S6 - Recall results**

| Sampling group | Eligible notifications | Notifications reviewed | False negatives (FN) | FN proportion, % (95% CI) | Estimated FN (95% CI) |
|---|---|---|---|---|---|
| **A** | 19 | 19 | 6 | 6.0 (0.3–0.3) | 6.0 (6.0–6.0) |
| **B1** | 84 | 23 | 23 | 84.0 (0.8–1.0) | 84.0 (69.8–84.0) |
| **B2** | 243 | 67 | 34 | 123.3 (0.4–0.6) | 123.3 (94.9–151.5) |
| **B3** | 468 | 130 | 66 | 237.6 (0.4–0.6) | 237.6 (197.9–277.1) |
| **C1** | 155 | 32 | 3 | 14.5 (0.0–0.2) | 14.5 (3.8–38.7) |
| **C2** | 8,304 | 135 | 1 | 61.5 (0.0–0.0) | 61.5 (0.0–372.9) |
| **C3** | 3,834 | 52 | 0 | 0.0 (0.0–0.1) | 0.0 (0.0–315.1) |
| **Total** | **13,107** | **458** | **133** | — | **527.0 (372.4–1,245.5)** |

Note: The expanded, sample-based assessment estimated overall linkage recall at approximately 98.1% (95% CI 95.4%-98.6%).

**Supplementary Table S7 - NHS number availability (A) and use of NHS-number-based match keys (B) by notification year, England, 2001–2024.**

| Notification year | Total notifications | NHS number available, n (%) | Total linked notifications | Linked via NHS-based MKs, n (%) |
|---|---|---|---|---|
| 2001 | 6,171 | 4005 (64.9) | 1,726 | 1411 (81.7) |
| 2002 | 6,674 | 4669 (70.0) | 1,763 | 1470 (83.4) |
| 2003 | 6,630 | 4794 (72.3) | 1,643 | 1395 (84.9) |
| 2004 | 6,926 | 5041 (72.8) | 1,639 | 1386 (84.6) |
| 2005 | 7,658 | 5711 (74.6) | 1,684 | 1468 (87.2) |
| 2006 | 7,684 | 5911 (76.9) | 1,618 | 1414 (87.4) |
| 2007 | 7,577 | 5953 (78.6) | 1,546 | 1375 (88.9) |
| 2008 | 7,809 | 6301 (80.7) | 1,474 | 1326 (90.0) |
| 2009 | 8,111 | 7002 (86.3) | 1,589 | 1484 (93.4) |
| 2010 | 7,674 | 6804 (88.7) | 1,452 | 1383 (95.2) |
| 2011 | 8,282 | 7487 (90.4) | 1,459 | 1406 (96.4) |
| 2012 | 8,087 | 7480 (92.5) | 1,376 | 1327 (96.4) |
| 2013 | 7,267 | 6827 (93.9) | 1,202 | 1163 (96.8) |
| 2014 | 6,474 | 6105 (94.3) | 1,117 | 1092 (97.8) |
| 2015 | 5,735 | 5468 (95.3) | 949 | 924 (97.4) |
| 2016 | 5,622 | 5389 (95.9) | 895 | 877 (98.0) |
| 2017 | 5,065 | 4870 (96.2) | 771 | 758 (98.3) |
| 2018 | 4,609 | 4480 (97.2) | 625 | 619 (99.0) |
| 2019 | 4,702 | 4543 (96.6) | 556 | 550 (98.9) |
| 2020 | 4,123 | 4022 (97.6) | 488 | 485 (99.4) |
| 2021 | 4,395 | 4305 (98.0) | 421 | 414 (98.3) |
| 2022 | 4,369 | 4312 (98.7) | 354 | 352 (99.4) |
| 2023 | 4,831 | 4768 (98.7) | 335 | 334 (99.7) |
| 2024 | 5,490 | 5429 (98.9) | 260 | 258 (99.2) |

**Supplementary Table S8 - NHS number availability by demographic and social characteristics among TB notifications, England, 2001–2024.**

| Characteristic | Category | Total notifications, n | NHS number absent, n (%) |
|---|---|---|---|
| **Age group** | 0–14 | 6,932 | 1,086 (15.7%) |
| | 15–44 | 90,369 | 13,479 (14.9%) |
| | 45–64 | 33,127 | 3,370 (10.2%) |
| | 65+ | 21,448 | 2,276 (10.6%) |
| | Missing | 89 | 78 (87.6%) |
| **Sex** | Male | 86,438 | 12,281 (14.2%) |
| | Female | 65,516 | 7,997 (12.2%) |
| | Missing | 11 | 11 (100.0%) |
| **UK-born** | Yes | 37,595 | 2,863 (7.6%) |
| | No | 106,885 | 15,822 (14.8%) |
| | Missing | 7,485 | 1,604 (21.4%) |
| **Ethnic group** | White | 32,028 | 2,652 (8.3%) |
| | Black | 34,316 | 5,931 (17.3%) |
| | South Asian | 62,463 | 7,635 (12.2%) |
| | Mixed/Other (incl. Chinese) | 19,066 | 3,193 (16.7%) |
| | Missing | 4,092 | 878 (21.5%) |
| **History of alcohol misuse** | Yes | 3,453 | 252 (7.3%) |
| | No | 83,761 | 4,531 (5.4%) |
| | Missing | 64,751 | 15,506 (23.9%) |
| **History of drug misuse** | Yes | 3,466 | 182 (5.3%) |
| | No | 84,553 | 4,789 (5.7%) |
| | Missing | 63,946 | 15,318 (24.0%) |
| **History of homelessness** | Yes | 3,938 | 504 (12.8%) |
| | No | 84,586 | 4,654 (5.5%) |
| | Missing | 63,441 | 15,131 (23.9%) |
| **History of imprisonment** | Yes | 3,283 | 346 (10.5%) |
| | No | 82,703 | 4,680 (5.7%) |
| | Missing | 65,979 | 15,263 (23.1%) |
| **Asylum seeker** | Yes | 1,747 | 172 (9.8%) |
| | No | 49,172 | 3,021 (6.1%) |
| | Missing | 101,046 | 17,096 (16.9%) |

*Note: Values are presented as n (row %). Percentages represent the proportion of notifications without an NHS number within each characteristic category. Missing values are included as a separate category.*

#### Supplementary Result 5. Sensitivity Analysis of PDS enrichment

Among 35,072 notifications with no NHS number before enrichment, 25,492 (72.7%) remained unlinked in both runs and 7,877 (22.5%) linked to the same ONS record. A further 441 (1.3%) linked only following enrichment, all through NHS-number-based keys: 382 through MK1 and 59 through MK2. Conversely, 1,158 (3.3%) linked only with pre-enrichment identifiers; 1,046 (90.3%) of these links arose through MK11-MK12, which had poor precision in the main validation.

Among 7,981 notifications linked in both runs, 7,877 (98.7%) linked to the same ONS record and 104 (1.3%) linked to different ONS records. All 104 changed from a demographic key (MK5-MK12) before enrichment to an NHS-number-based key (MK1-MK2) after enrichment; 73 (70.2%) had linked through MK11-MK12 before enrichment.

**Supplementary Table S9 - Sensitivity analyses of PDS NHS number enrichment**

| Linkage outcome | n | % |
|---|---|---|
| **A. Notifications with NHS number added through PDS enrichment (n=35,072)** | | |
| Unlinked in both runs | 25,492 | 72.7 |
| Linked to same ONS record in both runs | 7,877 | 22.5 |
| Linked post-PDS only | 441 | 1.3 |
| Linked pre-PDS only | 1,158 | 3.3 |
| Linked in both runs, but to different ONS records | 104 | 0.3 |
| **Total** | **35,072** | **100.0** |
| **B. Notifications with discordant original and PDS-derived NHS numbers (n=685)** | | |
| Unlinked using either NHS number | 531 | 77.5 |
| Linked using PDS-derived NHS number only | 149 | 21.8 |
| Linked using original NHS number only | 0 | 0.0 |
| Linked using both, but to different ONS records | 5 | 0.7 |
| **Total** | **685** | **100.0** |

Among the 685 notifications whose original and PDS-derived NHS numbers differed, 531 (77.5%) remained unlinked with either number. A further 149 (21.8%) linked only when the PDS-derived number was used, while five (0.7%) linked to different ONS records depending on which NHS number was used. No notification linked only when the original NHS number was used.

Overall, enrichment increased access to NHS-number-based match keys that demonstrated high precision and reduced reliance on later demographic keys. However, PDS-derived identifiers could not be independently verified within this analysis; the small set of discordant outcomes shows that upstream enrichment can also alter the mortality record selected. The sensitivity findings therefore support PDS enrichment as a linkage strength while retaining a caveat about possible propagated identifier error.

### Supplementary Result 6. Comparison of death records across NTBS, PDS and ONS

Agreement in recorded death status between NTBS, PDS and ONS was incomplete, reflecting differences in the purpose and coverage of the three systems (Figure S1). ONS mortality data are derived from formally registered deaths, whereas death information in PDS is recorded for demographic record management (refs). NTBS is a clinical surveillance system rather than a mortality register, therefore deaths may be recorded during routine TB case management, including for individuals diagnosed post-mortem or who die during treatment, while deaths occurring after follow-up are not routinely captured.

Of 3,470 deaths identified in ONS but not PDS, 2,400 (69.2%) occurred among individuals who had not matched to PDS during the PDS-enrichment process. Among notifications who had a PDS match, the absence of a PDS death record was not unexpected given the different purposes of the two systems.

Conversely, 799 notifications had a death recorded in PDS but no corresponding ONS mortality record identified through the linkage process. An exploratory review of five randomly selected PDS-only deaths on NHS Spine by a TB physician found that all five were recorded as informal deaths. NHS England confirmed that informal deaths may be entered manually by NHS organisations following notification of a patient's death and do not require a corresponding civil death registration. This may include deaths occurring overseas or deaths not registered through the General Register Office (25,31).

A further 268 individuals had a death recorded in NTBS but not identified in PDS or ONS, of whom 216 (80.6%) had not linked to PDS. NTBS-only deaths occurred throughout the study period, with no clear temporal pattern. Potential explanations include deaths occurring outside England and Wales or failure to identify corresponding records during linkage.

These findings indicate that discrepancies between NTBS, PDS and ONS largely reflect the differing purposes and mechanisms of the three systems, rather than deficiencies in the linkage process

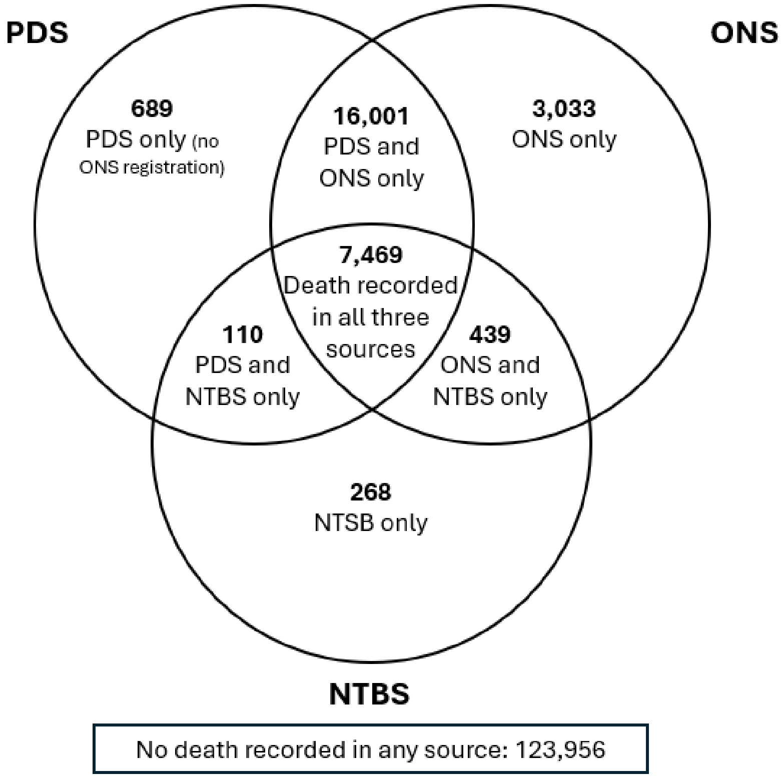

**Figure S1 - Death ascertainment across PDS, ONS and NTBS notifications in England, 2001-2024**
